# Impact of Differential Vaccine Effectiveness on COVID-19 Hospitalization Cases: Projections for 10 Developed Countries where Booster Vaccines were Recommended

**DOI:** 10.1101/2022.09.26.22280377

**Authors:** Michael Maschio, Kelly Fust, Amy Lee, Nicolas Van de Velde, Philip O. Buck, Michele A. Kohli

**Affiliations:** Quadrant Health Economics Inc; 92 Cottonwood Crescent, Cambridge, Ontario, Canada; Moderna Inc; 200 Technology Square, Cambridge, MA, United States

## Abstract

**Background & Objectives:** In a previous analysis, a decision-analytic model was used to analyze the clinical and economic impact of the differences in effectiveness between the two licensed mRNA COVID-19 booster vaccines, mRNA-1273 and BNT162b2, in 2022 for adults aged 18 years and older in the United States (US). In this analysis, the same model was used to estimate the impact that administering first booster doses with mRNA-1273 could have had on COVID-related hospitalizations and costs over a 6-month period in 10 developed countries (Australia, Canada, France, Germany, Italy, Japan, South Korea, Spain, United Kingdom [UK], and US), considering updated effectiveness data.

**Methods:** The model was used to estimate number of hospitalizations and related costs using the actual vaccine distribution for the first COVID-19 booster from each country. These estimates were compared to a scenario where 100% of doses for that 6-month period was assumed to be mRNA-1273. The effectiveness of mRNA-1273 compared to BNT162b2 was estimated from real world data from the UK.

**Results:** The total number of doses switched to the mRNA-1273 booster would range from 4.3 million in Spain to 39.4 million in Japan. The number of hospitalizations and associated hospitalization costs would be expected to fall in all countries, with the proportional decrease ranging from 1.1% (16,800 fewer) in Germany to 8.8% (25,100 fewer) in Australia.

**Conclusions:** Real-world effectiveness data suggest that a booster dose of the mRNA-1273 vaccine may be more effective compared to other vaccines used for booster doses. Given this difference in effectiveness, results of this analysis demonstrate that switching to 100% mRNA-1273 boosters would have reduced the number of hospitalizations and associated costs in each country during the first 6 months of the omicron period.

## INTRODUCTION

There are currently multiple vaccines used globally for active immunization to prevent coronavirus disease (COVID-19), including 2-dose messenger ribonucleic acid (mRNA) vaccines BNT162b2 (COMIRNATY®; Pfizer Inc, New York, NY, USA; BioNTech Manufacturing GmbH, Mainz, Germany) and mRNA-1273 (SPIKEVAX®; Moderna Inc, Cambridge, MA, USA). Other available vaccines include two viral vector-based vaccines, (single dose Ad26.COV2.S vaccine [Jcovden®; Johnson & Johnson, Beerse, Belgium] and 2-dose ChAdOx1-S vaccine [Vaxzevria®; AstraZeneca, Cambridge, England]), and the recently authorized 2-dose NVX-CoV2373 (Nuvaxovid® and Covovax®; COVID-19 Vaccine; Novavax Inc., Gaithersburg, MD, USA), a protein subunit vaccine. Booster doses are now recommended in many countries. Although the guidelines and recommendations for use of each vaccine vary by country, many countries specify a preference for mRNA vaccines for booster doses, citing effectiveness and safety data.^1,2^

In a previously conducted analysis, Maschio et al.^3^ examined the clinical and economic impact of the differences in effectiveness between mRNA-1273 and BNT162b2 booster vaccinations over one year (2022) in United States (US) adults aged 18 years and older. At the time that the original analysis was completed, there were data for vaccine effectiveness (VE) against Delta but very few data on VE against Omicron. Therefore, Maschio et al. relied on estimates of VE from studies of neutralizing antibody titers or from small studies using one vaccine only. To utilize real world data, relative VE ratios between Omicron and Delta variants for BNT162b2 were applied to all other vaccines. In the current analysis, the most recent evidence for VE is used to estimate the impact of COVID-19 boosters on Omicron infections.

Real world evidence of the VE against Omicron are now available from studies with large sample sizes and meta-analyses.^4,5,6,7^ Butt et al.^5^ used a retrospective cohort matched study design to assess the VE of an mRNA-based booster dose compared to the primary series among members of the Veteran’s Affairs healthcare system (median age 72 years), who are at high risk of infection and adverse outcomes from COVID-19. Ono et al.^7^ conducted a study of comparative effectiveness between Japanese residents aged ≥16 years who received the BNT162b2 primary series and either a mRNA-1273 or BNT162b2 booster dose; due to data availability, they were unable to adjust for known risk factors for COVID-19. EPI-PHARE^6^ conducted a cohort study in adults ≥18 years to assess first booster dose efficacy (BNT162b2 or mRNA-1273) against risk of hospitalization for COVID-19 in France. Hulme et al.^4^ conducted a matched database analysis to compare the effectiveness of boosting adults aged ≥18 years in England with either BNT162b2 or mRNA-1273. As this study includes a large sample of the general adult population and directly compares the effectiveness of BNT162b2 and mRNA-1273 against COVID-19 hospitalization and death while controlling for potential differences in vaccine recipients, it provided the data required to update the previous analysis.

The current modelled analysis expands on the Maschio et al. study^3^ by adapting the previously developed decision analytic model to 9 additional countries for the period in which Omicron was the dominant variant of concern. The objective of this study was to estimate the impact that administering all first booster doses with mRNA-1273 could have had on COVID-related hospitalizations and costs over the 6-month Omicron period in the following countries that preferentially recommend the use of mRNA vaccines for boosters: Australia, Canada, France, Germany, Italy, Japan, South Korea, Spain, United Kingdom (UK), and US.

## METHODS

A previously described decision analytic model^3^ was used to compare two mRNA COVID-19 booster scenarios and estimate the potential number of hospitalizations and associated costs for each scenario by country. The target population for all countries was adults aged 18 years or older, with the exception of France, where policy limits mRNA-1273 to adults aged 30 years or older.^8^ The months of the analytic time frame were selected for consistency with when Omicron became the dominant variant of concern in each country. The time horizon for all analyses in the present study was the 6 month period from January to June 2022, with the exception of the UK and US, which were analyzed over the 6 month period from December 2021 to May 2022.

To construct the two scenarios for each country, the booster coverage rate and the proportion of different booster doses delivered were determined. The current scenario reflects the actual historical data on types of doses given as shown in Table 1. As mRNA vaccines are preferentially recommended in these countries, only a small portion of people (1.5% in the US; <1% in other countries) received another type of vaccine (ChAdOx1-S, Ad26.COV2.S, or NVX-CoV2373). In the second mRNA-1273 scenario, the booster coverage rates are identical to the current scenario, but all individuals received the mRNA-1273 booster.

**Table 1.** Model inputs: Proportions of different vaccine types delivered for the first booster for each country.

|  | mRNA-1273 | BNT162b2 | Other (non-mRNA) |
| --- | --- | --- | --- |
| **Australia**<sup>9</sup> | 12.0% | 87.6% | 0.4% |
| **Canada**<sup>10</sup> | 43.6% | 56.1% | 0.3% |
| **France**<sup>10</sup> | 30.0% | 70.0% | 0.0% |
| **Germany**<sup>51</sup> | 36.3% | 63.7% | 0.0% |
| **Italy**<sup>18</sup> |  |  |  |
| Ages 18-59 | 55.1% | 44.9% | <0.1% |
| Ages ≥60 | 41.7% | 58.3% | <0.1% |
| **Japan**<sup>49</sup> | 42.3% | 57.7% | <0.1% |
| **South Korea**<sup>44</sup> | 37.6% | 62.3% | 0.2% |
| **Spain**<sup>11</sup> | 54.0% | 46.0% | 0.0% |
| **United Kingdom**<sup>12</sup> | 23.4% | 76.4% | 0.2% |
| **United States**<sup>13,14</sup> | 42.6% | 55.9% | 1.5% |

Row totals may not sum to 100% due to rounding.

In the model, the baseline risk of infection in the unvaccinated varies by month. Amongst those who received the primary series only or the primary series plus booster, the incidence of infection is reduced according to VE, and is recalculated each month to account for waning based on time since vaccination. The coverage and mix of primary series vaccines delivered is the same for the current scenario and the all mRNA-1273 scenario. For those who received the primary series in 2021, we made the simplifying assumption that their series was completed in the month displayed in Appendix Table 7, as this marked the month at which 50% cumulative coverage in the population was achieved in each country. This month, which varies by country, was therefore our anchor month to calculate time since primary series completion for the VE calculations for each month. The primary series VE was estimated as an average of the waning adjusted VE weighted by the proportion of the cohort receiving each primary series. For anyone who had received a booster prior to the start of the analytical time horizon, we made the simplifying assumption that the booster was received in the month prior. As people continued to receive boosters in 2022, the booster VE was estimated as a weighted average of the waning adjusted VE (time since receiving their booster) and the proportion of the cohort receiving each booster.

### Model Inputs: Primary Series Vaccine Effectiveness

Primary series VE is summarized in Table 2. Primary series VE for all vaccines against infection and monthly waning for mRNA-1273 and BNT162b2 were obtained from a meta-analysis conducted by Pratama et al., 2022.^15^ Pratama identified studies available by April 6, 2022, including 12 from the US, 4 from Europe, 2 from South Africa, 2 from Qatar and 1 from Canada. Monthly waning for Ad26.COV2.S and ChAdOX1-S were calculated from a large study by Adams et al., 2022 in the US,^16^ and the UK Health Security Agency (HSA) COVID-19 report, respectively.^17^

**Table 2.** Model Inputs: Primary Series and First Booster Effectiveness^15,16,17,4^.

|  | Primary Series |  | Booster |  |
| --- | --- | --- | --- | --- |
|  | Initial Effectiveness | Monthly Waning | Initial Effectiveness | Monthly Waning |
| **Infection** |  |  |  |  |
| mRNA-1273 | 76.1% | 9.6% | 71.3% | 5.4% |
| BNT162b2 | 58.4% | 5.9% | 68.8% | 5.9% |
| Ad26.COV2.S | 54.0% | 5.3% | 69.3% | 5.3% |
| ChAdOx1-S | 56.0% | 7.2% | 69.3% | 5.3% |
| **Hospitalization** |  |  |  |  |
| mRNA-1273 | 90.0% | 3.6% | 92.7% | 1.8% |
| BNT162b2 | 89.9% | 2.7% | 89.1% | 2.7% |
| Ad26.COV2.S | 67.0% | 5.3% | 86.0% | 5.3% |
| ChAdOx1-S | 85.0% | 4.4% | 86.0% | 5.3% |

Primary series hospitalization VE from BNT162b2 and Ad26.COV2.S, as well as monthly waning for BNT162b2, were obtained from Pratama et al., 2022.^15^ Data for mRNA-1273 and ChAdOx1-S were not available from Pratama, and therefore VE against hospitalization and waning were calculated from the UK HSA COVID-19 report.^17^ Waning data for Ad26.COV2.S were unavailable and assumed to be the same as waning against infection.

The average VE was calculated by country as a weighted average using the proportion of doses delivered shown in Table 3 as weights. In all countries, the most common primary series received was a mRNA vaccine. The UK provided the highest proportion of ChAdOX1-S. Overall, the proportion of people receiving Ad26.COV2.S was low. As NVX-CoV2373 was only recently approved, there are no VE data available for the Omicron variant. As less than 0.1% of patients received it in only a few of the countries of interest, it was not included in the analysis.

### Model Inputs: Booster Vaccines Effectiveness

Booster VEs against infection for BNT162b2 and ChAdOx1-S were obtained from Pratama et al., 2022.^15^ As the current analysis focuses on the difference in booster VE of the two mRNA vaccines, it was important to capture the true relative difference between the two. In order to leverage the real-world evidence data published by Hulme et al., 2022, the hazard ratio (HR) comparing the VE of mRNA-1273 to BNT162b2 was applied to the VE of BNT162b2 to obtain the VE of mRNA-1273. Given the small proportion of doses of Ad26.COV2.S delivered globally as a booster, data were unavailable, and were approximated by calculating the ratio between Ad26.COV2.S primary series infection to hospitalization VEs and applying it to Ad26.COV2.S booster hospitalization VE.

Booster VE against hospitalization for BNT162b2 and Ad26.COV2.S were obtained from Pratama et al., 2022.^15^ Similar to booster VE against infection, the HR from Hulme et al., 2022^4^ for hospitalizations was applied the BNT162b2 booster hospitalization VE to obtain the VE of mRNA-1273. Data for ChAdOx1-S were unavailable from the Pratama meta-analysis. In the US, only 1.5% of boosters delivered were not an mRNA vaccine and for the other countries, this proportion was less than 0.5% (See Table 1). Data on the performance of these boosters, especially ChAdOx1-S are sparse, and we therefore assigned them all the infection and hospitalization effectiveness associated with Ad26.COV2.S which represented the largest proportion of the non-mRNA vaccines (Table 2).

Booster waning rates for infection and hospitalization were assumed to be the same as primary series for all vaccines except for mRNA-1273. As the purpose of the analysis was to estimate the impact of the difference in VE noted by Hulme et al., 2022, the booster waning rates for mRNA-1273 were adjusted to maintain the HRs from this study for the entire 6-month duration.^4^ Sensitivity analyses on the effectiveness of mRNA-1273 were performed by varying the HRs estimated by Hulme et al. using the 95% confidence intervals (CI). Using the lower bound yields estimates of VE against infection and hospitalization of 71.6% (waning 5.4%) and 93.6% (waning 1.58%), respectively. Using the upper bound yields estimates of VE against infection and hospitalization of 71.3% (waning 5.4%) and 91.5% (waning 2.12%), respectively.

### Other Model Inputs

Model inputs were estimated for each country from publicly available data and other published sources as available. Estimates of the total population size in each country were based on United Nations data^19^ (Appendix Table 6). Primary series^44-53^ and first booster coverage rates^44-53^ were also estimated for each country, by age group as available (Appendix Table 7 and Figure 3 to Figure 12). Hospitalization rates, by age group as available, and costs were estimated based on local data for each country (Table 4).^22 -43^ Sensitivity analyses were performed by varying hospitalization rates ±25%. Hospitalization costs represent the average cost per hospital stay for COVID-19 infection. Similarly to the previously published analysis,^3^ infection incidence estimates amongst the unvaccinated in each country over the 6-month time horizon were based on modeling from the Institute for Health Metrics and Evaluation (IHME) (Figure 1), although the incidence rates used (updated July 19, 2022) do differ from the past projections for this period.^20^,21

**Table 3.** Model Inputs: Primary Series Vaccine Distribution.

|  | **mRNA-1273** | **BNT162b2** | **Ad26.COV2.S** | **ChAdOx1-S\*** |
| --- | --- | --- | --- | --- |
| **Australia**<sup>9</sup> | 0.9% | 79.9% | 0.0% | 19.2% |
| **Canada**<sup>10</sup> | 21.2% | 76.0% | 0.2% | 2.6% |
| **France**<sup>10</sup> | 12.0% | 82.0% | 0.0% | 6.0% |
| **Germany**<sup>51</sup> | 10.4% | 83.7% | 0.0% | 5.8% |
| **Italy**<sup>18</sup> |  |  |  |  |
| Ages 18-59 | 16.7% | 75.2% | 1.6% | 6.4% |
| Ages ≥60 | 10.9% | 61.7% | 1.8% | 25.5% |
| **Japan**<sup>49</sup> | 16.7% | 83.3% | 0.0% | 0.1% |
| **South Korea**<sup>44</sup> | 15.3% | 59.3% | 3.6% | 21.9% |
| **Spain**<sup>11</sup> | 13.0% | 70.0% | 5.0% | 12.0% |
| **United Kingdom**<sup>12</sup> | 3.2% | 49.8% | 0.0% | 47.1% |
| **United States**<sup>13,14</sup> | 34.8% | 57.5% | 7.7% | 0.0% |

\*Includes NVX-CoV2373 in South Korea, Japan, Germany, and Australia, but proportion is less than 1%. Row totals may not sum to 100% due to rounding.

**Table 4.** Model Inputs: Hospitalization Rates in Unvaccinated Individuals and Hospitalization Costs.

|  | **Hospitalization Rates, by age** | **Cost per Hospitalization** |
| --- | --- | --- |
| **Australia**<sup>22,46, 23</sup> | 18-29 years: 5.6%<br>30-39 years: 9.2%<br>40-49 years: 11.6%<br>50-59 years: 13.6%<br>60-69 years: 17.9%<br>70-79 years: 24.4%<br>80+ years: 30.9% | AUD 23,000 |
| **Canada**<sup>24,25</sup> | ≥18: 5.5% | CAD 25,103 |
| **France**<sup>26,27</sup> | 30+*: 11.7% | € 7,044 |
| **Germany**<sup>28,29,30</sup> | 18-59: 5.14%<br>≥60: 33% | € 15,394 |
| **Italy**<sup>31</sup> | 18-39: 1.85%<br>40-59: 6%<br>60-79: 20.3%<br>≥80: 36.7% | € 9,290 |
| **Japan**<sup>32,33</sup> | ≥18: 13.7% | ¥ 1,732,027 |
| **South Korea**<sup>34,35</sup> | ≥18: 8.3% | USD 4,350 |
| **Spain**<sup>36,37</sup> | 18-29: 1.5%<br>30-59: 5.1%<br>60-79: 18.2%<br>≥80: 36.0% | € 7,050 |
| **United Kingdom**<sup>38,39,40</sup> | 20-29: 0.90%<br>30-39: 2.40%<br>40-49: 3.68%<br>50-59: 7.65%<br>60-69: 12.45%<br>70-79: 18.23%<br>≥80: 20.5% | £ 3,661 |
| **United States**<sup>41,42,43</sup> | 18 - 49 years: 1.25%<br>50 - 64 years: 3.45%<br>65+ years: 15.71% | USD 19,460 |

\*Only those ages 30 years and above were considered for France as mRNA-1273 was not used in younger age groups.

**Figure 1.**
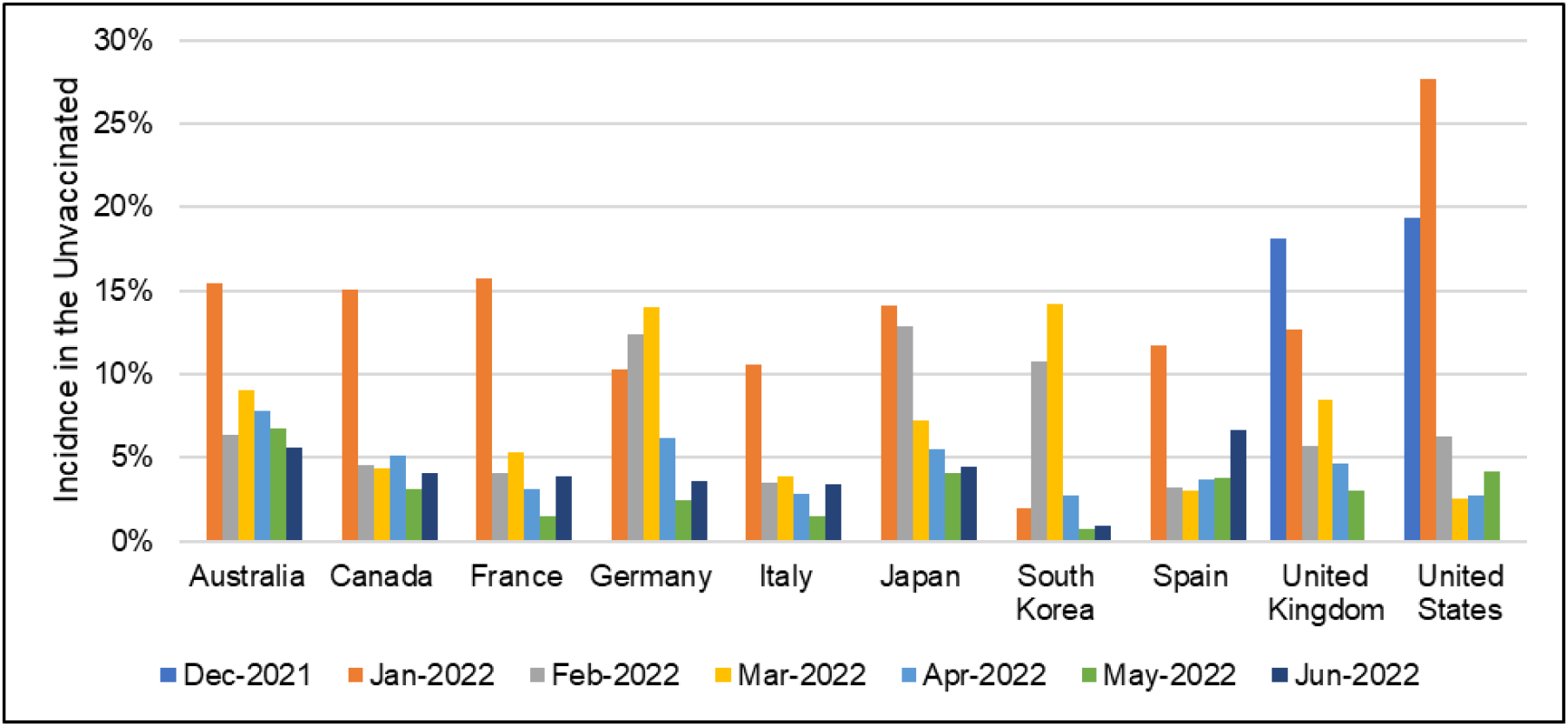
Model Inputs: Country-Specific Incidence of COVID-19 Rates in the Unvaccinated Population During the 6-Month Analytic Time Horizon^20,21^.

## RESULTS

The total number of hospitalizations and hospitalization costs for each first booster scenario are presented in Table 5, along with the total number of doses that hypothetically would be switched if mRNA-1273 was used exclusively for the booster dose. Based on the actual historical vaccine use for the booster doses, the model predicts the total numbers of hospitalizations for each country, ranging from 175,900 in Canada to 2,562,900 in the US. If 100% of boosted individuals had received the mRNA-1273 booster the total number of doses switched to the mRNA-1273 booster would range from 4.3 million in Spain to 39.4 million in Japan (Table 5). Given the higher overall effectiveness of mRNA-1273 relative to the BNT162b2 booster, the number of hospitalizations would be expected to fall in each country in the 100% mRNA-1273 scenario, with the proportional decrease in hospitalizations ranging from 1.1% (16,800 fewer) in Germany to 8.8% (25,100 fewer) in Australia. Given the observed reductions in the numbers of hospitalizations predicted by the model, hospitalization costs also fell in each country, paralleling the observed percentage reductions in the number of hospitalizations (Table 5). In sensitivity analyses varying hospitalization rates, the absolute number of hospitalizations increased or decreased but the percentage change in the number of hospitalizations for the mRNA-1273 scenario did not alter from the base-case because all parameters in the model are linearly related and therefore change proportionally. Results of the effectiveness sensitivity analysis (Figure 2) demonstrate that the impact of variation on the percentage change in hospitalizations is proportional to the percentage change observed in the base case. For example, in those countries with a higher percentage change in the number of hospitalizations between the current and mRNA-1273 scenarios, the impact of the variation in effectiveness was greater. In all sensitivity analyses, there was still a decrease in hospitalization for all scenarios where 100% of the population was switched to mRNA-1273.

**Figure 2.**
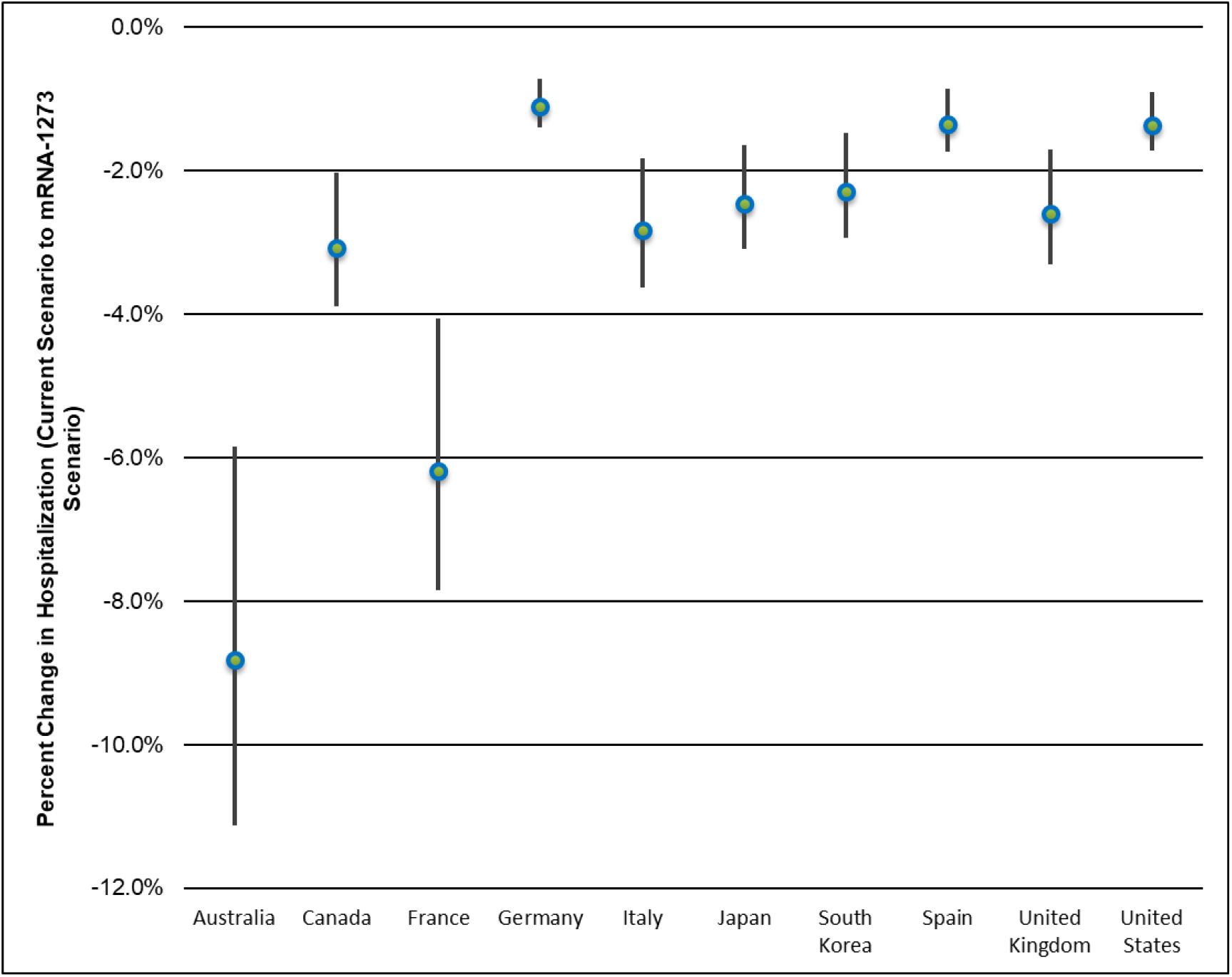
Impact of Effectiveness Sensitivity Analysis on the Percent Change in Hospitalizations.

**Table 5.** Base-Case Results.

|  | Historical Booster Vaccine Use (Current Scenario) |  |  | Change from Current Scenario to Exclusive Use of mRNA-1273 for Booster |  |  |  |
| --- | --- | --- | --- | --- | --- | --- | --- |
|  | Doses Administered (mil) | Total # of Hospitalizations | Total Hospitalization Costs (mil) | Doses (mil) Switched to mRNA-1273 | Change in # of Hospitalizations | Change in Hospitalization Costs (mil) | % Change in Hospitalizations and Associated Costs* |
| **Australia** | 11.0 | 284,800 | AUD 6,600 | 9.6 | -25,100 | -AUD 580 | -8.8% |
| **Canada** | 11.7 | 175,900 | CAD 4,400 | 6.6 | -5,400 | -CAD 130 | -3.1% |
| **France** | 20.6 | 369,000 | €2,600 | 14.4 | -22,800 | -€160 | -6.2% |
| **Germany** | 8.4 | 1,533,300 | €23,600 | 5.4 | -16,800 | -€ 260 | -1.1% |
| **Italy** | 18.2 | 369,500 | €3,400 | 9.0 | -10,500 | -€97 | -2.8% |
| **Japan** | 68.3 | 1,905,300 | ¥3,300,000 | 39.4 | -46,800 | -¥81,100 | -2.5% |
| **South Korea** | 10.5 | 373,300 | USD 1,600 | 6.6 | -8,600 | -USD 40 | -2.5% |
| **Spain** | 9.3 | 300,700 | €2,100 | 4.3 | -4,100 | -€29 | -1.3% |
| **United Kingdom** | 12.9 | 528,600 | £1,900 | 9.9 | -13,700 | -£50 | -2.6% |
| **United States** | 53.0 | 2,562,900 | USD 49,900 | 30.4 | -34,900 | -USD 690 | -1.4% |

Mil – million; # number; % percent
\*The percent change in hospitalizations with the change to 100% mRNA-1273 doses is the same as the percent change in costs associated with hospitalizations

## DISCUSSION

This study found that, when compared to the current use of multiple COVD-19 vaccines for booster doses in the 10 countries assessed, shifting to 100% mRNA-1273 boosters would have decreased the expected number of hospitalizations, which in turn would have reduced COVID-19 related hospitalization costs. In the current scenario, a portion of people received non-mRNA vaccines as boosters but as this is only 1.5% in the US and less than 1% in other countries, these have minimal impact on results. Results of the effectiveness sensitivity analysis demonstrate that the impact of variation on the percentage change in hospitalizations depends on the observed percentage change in the base-case analysis. Furthermore, sensitivity analyses varying hospitalization rates resulted in changes to the absolute number of hospitalizations but did not alter the percentage change in the number of hospitalizations for the mRNA-1273 scenario. In all scenarios, switching to 100% mRNA-1273 was predicted to decrease hospitalization. These findings are subject to several limitations discussed below, some of which have been previously described in Maschio et al.^3^

There is high uncertainty regarding VE against emerging variants. This study aimed to build upon Maschio et al. by using historical data from the Omicron period, as well as incorporating recently published data into effectiveness estimates. Although the effectiveness data represent a mixture of the Delta and Omicron variants, Hulme et al.^4^ is one of the few publications that presents a controlled comparison of mRNA-1273 and BNT162b2 in the general population during the Omicron wave. Results from Hulme et al. are consistent with other studies published during the Delta and Omicron periods.^5,6,7^ However, these alternative studies do not control for bias in the same manner or have smaller sample sizes. Additionally, the incidence data utilized in this analysis represent the rapid rise and subsequent decline in cases amid the Omicron wave. A lack of COVID-19 testing during the Omicron wave may have led to an underestimation of true infection rates, although IHME numbers include corrections to account for this issue. A sharp increase in incidence occurred across all countries during the time horizon of the analysis and this may not be observed again in the future. Conversely, it is unknown whether a future variant of concern may be more virulent than Omicron. As such, it is difficult to use the findings from the present analysis to inform predictions about the ability of mRNA-1273 to reduce the hospitalization burden in the future; rather, the findings from this study should be viewed through a historical lens.

Although the present analysis is focused on the Omicron period, in the real-world, first booster doses were delivered prior to the start of the analytic time frame. In the model, first booster doses delivered prior to the start of the analyses (December 2021 or January 2022) were not eligible for switching to mRNA-1273. As such, the historical impact of switching first booster doses to mRNA-1273 may be underestimated. Finally, the quality of input data vary by country. For example, age-specific coverage rates and hospitalization data were not available for all settings. Age is a known driver of COVID-19 vaccination uptake and hospitalization risk; as such, a lack of age-specific data may lead to an over-or underestimation of the impact of mRNA-1273 in these countries.

## CONCLUSIONS

Real-world effectiveness data suggest that a booster dose of the mRNA-1273 vaccine may be more effective compared to other vaccines used for booster doses. Given this difference in effectiveness, results of this analysis demonstrate that switching to 100% mRNA-1273 boosters would have reduced the number of hospitalizations and associated costs in each country during the first 6 months of the omicron period.

## Data Availability

All data produced in the present work are contained in the manuscript.

## APPENTIX

**Table 6.** Population Size, by Country and Age Group.

| Population, by Age Group (million)<sup>19</sup> |  |  |  |  |  |  |  |  |
| --- | --- | --- | --- | --- | --- | --- | --- | --- |
| South Korea | Additional age groups not included |  |  |  |  |  |  | **Total (≥18)** |
|  |  |  |  |  |  |  |  | 44.5 |
| Canada | **18-59** | **≥60** | Additional age groups not included |  |  |  |  | **Total** |
|  | 21.3 | 10.0 |  |  |  |  |  | 31.2 |
| Australia | **18-29** | **30-39** | **40-49** | **50-59** | **60-69** | **70-79** | **≥80** | **Total** |
|  | 4.8 | 3.9 | 3.3 | 3.2 | 2.8 | 2.0 | 1.1 | 21.1 |
| Italy | **18-24** | **25-39** | **40-49** | **50-59** | **60-69** | **70-79** | **≥80** | **Total** |
|  | 4.1 | 9.7 | 8.4 | 9.6 | 7.7 | 6.0 | 4.6 | 50.0 |
|  | **18-24** | **25-29** | **30-39** | **40-54** | **55-64** | **65-74** | **≥75** | **Total** |
| France | Not included |  | 7.8 | 12.5 | 8.2 | 7.4 | 6.6 | 42.4 |
| Japan | 8.3 | 6.1 | 13.2 | 26.3 | 15.2 | 16.9 | 20.2 | 106.3 |
| Spain | **18-24** | **25-29** | **30-49** | **50-59** | **60-69** | **70-79** | **≥80** | **Total** |
|  | 3.4 | 2.5 | 13.5 | 7.3 | 5.7 | 4.1 | 2.9 | 39.5 |
|  | **18-29** | **30-39** | **40-49** | **50-59** | **60-69** | **70-79** | **≥80** | **Total** |
| Germany | 11.1 | 10.9 | 10.1 | 13.3 | 10.7 | 7.4 | 5.9 | 69.4 |
| United Kingdom | 9.8 | 9.0 | 8.3 | 9.1 | 7.5 | 6.0 | 3.5 | 53.4 |
| United States | **18-24** | **25-39** | **40-49** | **50-64** | **65-74** | **75-84** | **≥85** | **Total** |
|  | 31.0 | 69.0 | 42.1 | 64.1 | 33.9 | 17.5 | 6.6 | 264.1 |

**Table 7.** Primary Series Coverage, by Country and Age Group.

| Primary Series Coverage, by Age Group* |  |  |  |  |  |  |  | 50% of Primary Coverage Achieved** |
| --- | --- | --- | --- | --- | --- | --- | --- | --- |
| South Korea<sup>44</sup> | ≥18 | Additional age groups not included |  |  |  |  |  | September 2021 |
|  | 82.3% |  |  |  |  |  |  |  |
| Canada<sup>45</sup> | 18-59 | ≥60 | Additional age groups not included |  |  |  |  | July 2021 |
|  | 85.2% | 93.6% |  |  |  |  |  |  |
| Australia<sup>46</sup> | 18-29 | 30-39 | 40-49 | 50-59 | 60-69 | 70-79 | ≥80 | October 2021 |
|  | 83.6% | 89.1% | 91.4% | 93.9% | 96.0% | 100.0% |  |  |
| Italy<sup>47</sup> | 18-24 | 25-39 | 40-49 | 50-59 | 60-69 | 70-79 | ≥80 | July 2021 |
|  | 82.7% | 79.2% |  | 83.7% | 89.3% |  |  |  |
|  | 18-24 | 25-29 | 30-39 | 40-54 | 55-64 | 65-74 | ≥75 |  |
| France<sup>48</sup> | Not included |  | 88.2% | 90.4% | 92.2% | 94.0% | 90.1% | July 2021 |
| Japan<sup>49,50</sup> | 85.1% | 85.5% | 86.8% | 89.4% | 91.8% | 94.0% | 88.3% | August 2021 |
| Spain<sup>47</sup> | 18-24 | 25-29 | 30-49 | 50-59 | 60-69 | 70-79 | ≥80 | July 2021 |
|  | 71.2% | 77.8% |  | 87.8% | 96.4% |  |  |  |
|  | 18-29 | 30-39 | 40-49 | 50-59 | 60-69 | 70-79 | ≥80 |  |
| Germany<sup>51</sup> | 74.0% | 74.9% | 74.1% | 73.1% | 87.1% | 84.4% | 86.6% | August 2021 |
| United Kingdom<sup>52</sup> | 56.1% | 63.1% | 74.4% | 84.8% | 89.4% | 93.9% | 94.7% | May 2021 |
| United States<sup>53</sup> | 18-24 | 25-39 | 40-49 | 50-64 | 65-74 | 75-84 | ≥85 | May 2021 |
|  | 58.5% | 62.6% | 71.0% | 78.1% | 90.0% | 84.6% |  |  |

\*Estimates are applied to the month prior to start of analysis (November 2021 for December 2021 – May 2022 analysis and December 2021 for January – June 2022 analysis)
\*\*Similar to Maschio et al., the analysis included the months at which 50% coverage of the primary series was reached to estimate waning. Primary series coverage of 50% was reached in May 2021 (UK, US), July 2021 (Canada, France, Italy, Spain), August 2021 (Germany, Japan), September 2021 (South Korea), and October 2021 (Australia).

**Figure 3.**
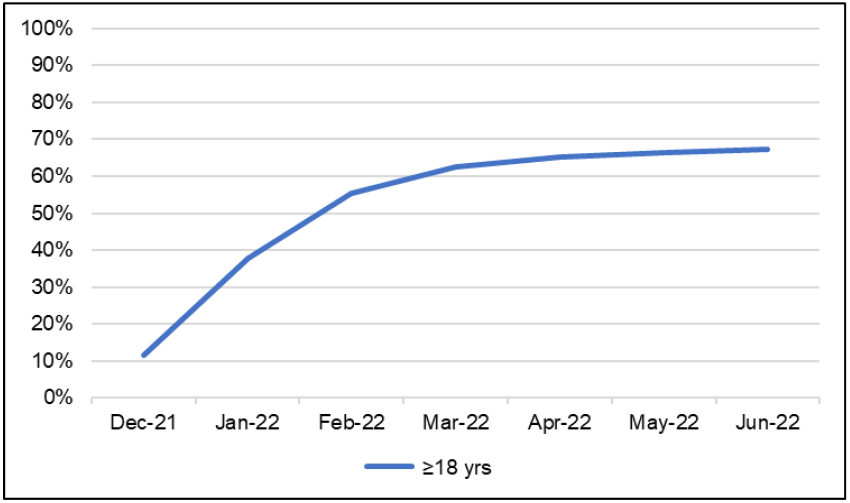
Australia First Booster Coverage^46^.

**Figure 4.**
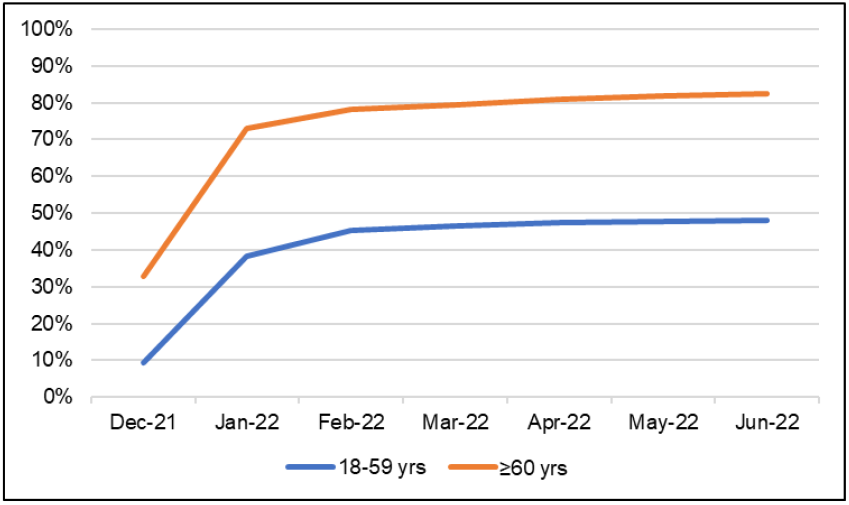
Canada First Booster Coverage^45^.

**Figure 5.**
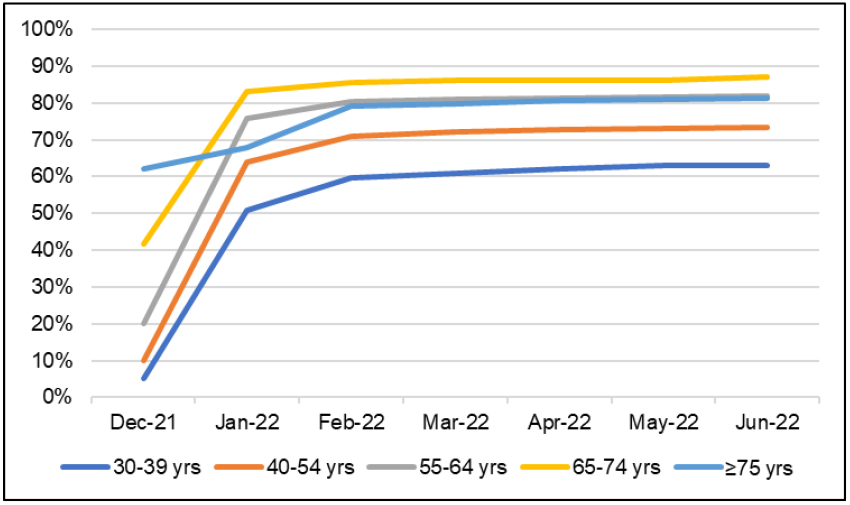
France First Booster Coverage^48^.

**Figure 6.**
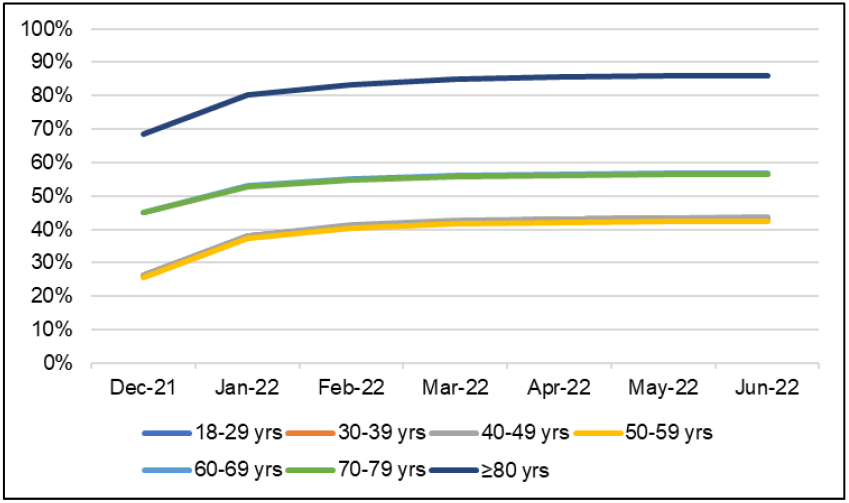
Germany First Booster Coverage^51^.

**Figure 7.**
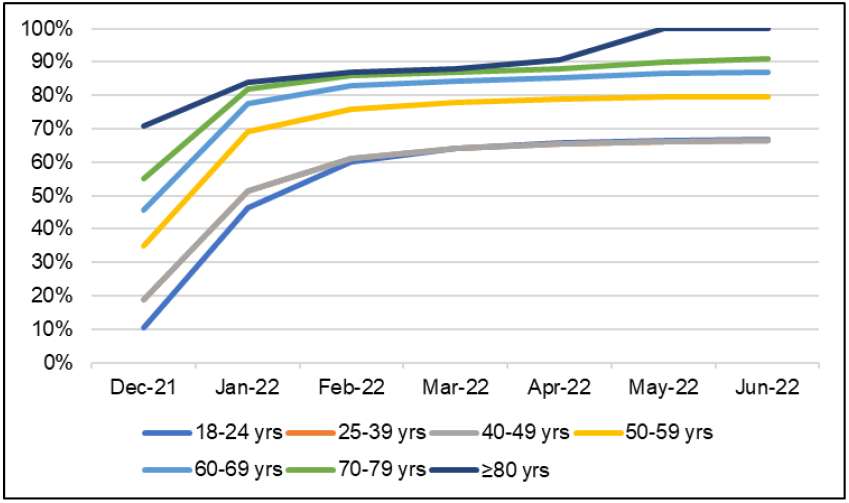
Italy First Booster Coverage^47^.

**Figure 8.**
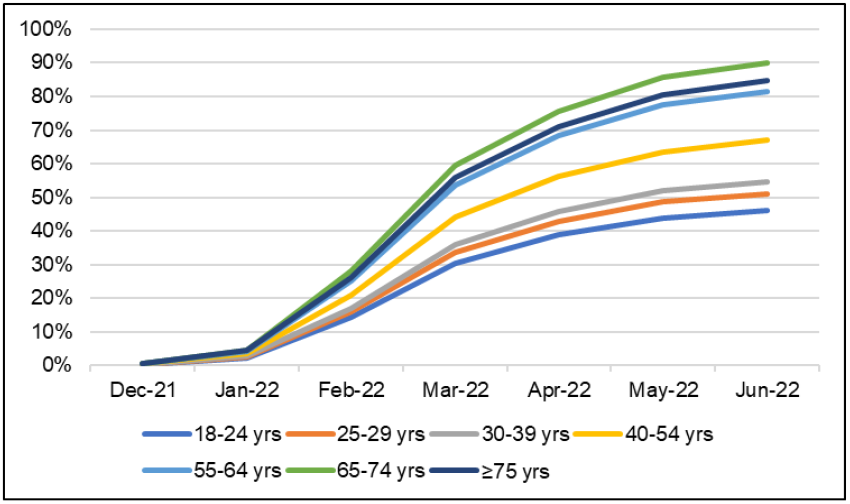
Japan First Booster Coverage^49,50^.

**Figure 9.**
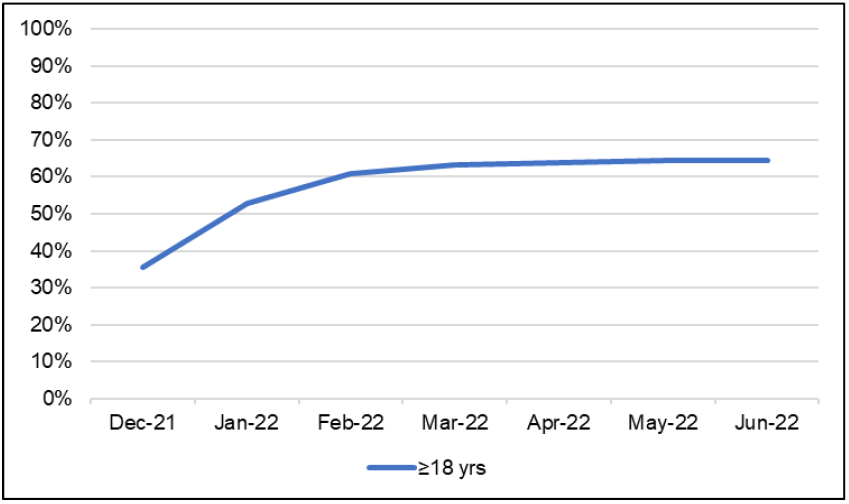
South Korea First Booster Coverage^44^.

**Figure 10.**
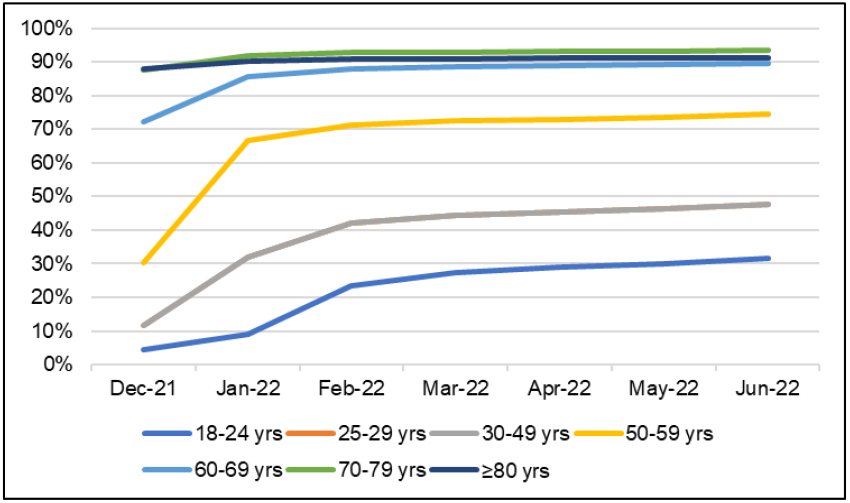
Spain First Booster Coverage^47^.

**Figure 11.**
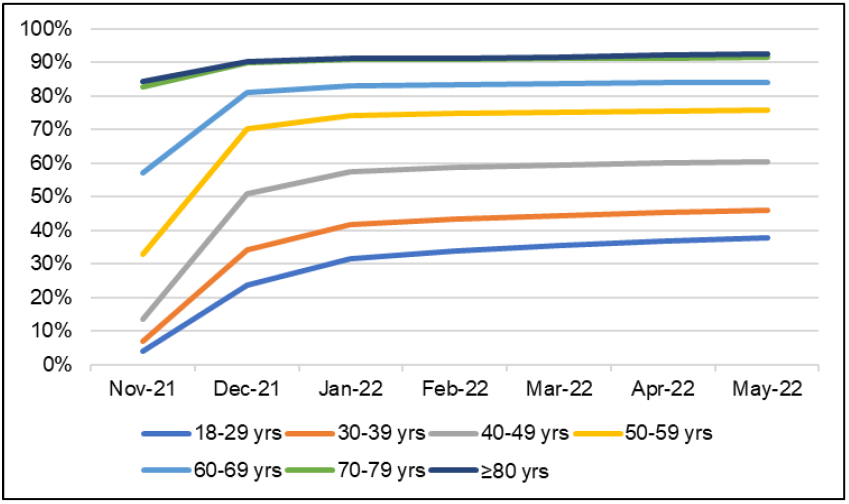
United Kingdom First Booster Coverage^52^.

**Figure 12.**
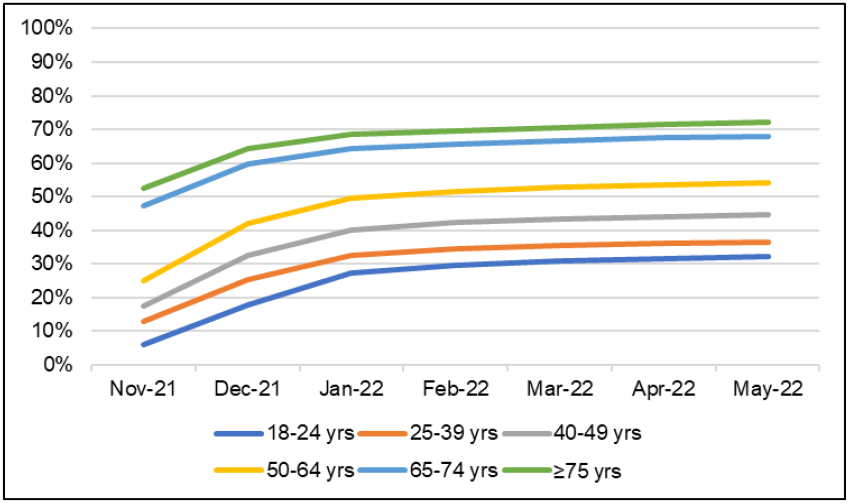
United States First Booster Coverage^53^.

All of the above figures show vaccine coverage calculated as the percent of vaccines delivered to the number of people in the age group.

